# Extended Hematology-Oncology Fellowships in Bone Marrow Transplantation, Leukemia, Lymphoma, and Myeloma: What Proportion of Trainees Train and Remain at these Centers?

**DOI:** 10.1101/2023.07.14.23292614

**Authors:** Vinay Prasad

## Abstract

**Introduction:** A number of extra fellowships after traditional hematology oncology training have started in recent years, including training in lymphoma, myeloma, bone marrow transplantation, and leukemia. The benefit of this additional training is disputed.

**Methods and Materials:** We sought to quantify, from publicly reported data, what percentage of trainees either performed the extra year at the same location as they performed their fellowship– raising the question why they did not gain this training during the regular fellowship years– and what fraction stayed on at the same institution–raising the question of why the training was not performed on the job. Google and LinkedIn searches were performed to assemble the dataset.

**Results:** We examined 6 programs that made their graduate information public. We found 30% (17/56) of fellows either trained at the same institution or remained at the same institution as their extra fellowship.

**Conclusion:** Nearly 1 in 3 trainees that forgo an extra year do so in a context where they had opportunity to work at that center before or afterwards, raising the question as to whether the extra year was needed or could have been built into existing professional obligations.

## Introduction

Physicians spend an average of 14 years preparing for their future role as an attending in the United States of America (US).^1^ As a time-intensive process with financial and health burdens for both physicians and patients, there have been many calls to reimagine the medical educational system.^2^ Fellowships represent the final training step on the path to becoming an independent, typically specialist, physician. Initially created to bolster expertise, medical fellowships aim to provide physicians with a more precise and deeper understanding of a specific area in medicine. The number of fellowships has grown over the last 5 decades. In the 1960s, there were a few dozen specialist boards, but in 2011, there were over 150.^3^

Recently, in the field of hematology oncology, there has been the growth of specialized additional fellowships in hematologic malignancies. These include fellowships in bone marrow transplant (BMT), myeloma, leukemia, and lymphoma. The validity and need of these fellowships has been debated.^4,5^ One argument in favor of additional fellowships is that individuals may have trained in hematology oncology programs that lack detailed instruction in these fields, or that they may wish to take these skills to new destinations and institutions.

As such, I sought to examine the characteristics of a systematic sample of hematology oncology fellows who completed these extra years of fellowship. Specifically, I asked what percent previously trained at those institutions– raising the question of why they did not gain this training during the regular fellowship years– and what fraction stayed with that institution–raising the question of why the training was not performed on the job.^6^

## Methods

I sought to assemble a dataset of physicians who performed extra years of training in hematologic malignancies. Specifically, I examined extra fellowships after hematology and medical oncology, including lymphoma, myeloma, leukemia, and bone marrow transplant.

In order to build the data set, a google search was performed for extra fellowships in each of these areas from March to May 2023. For each search, the first 5 pages of results were reviewed to identify programs for data abstraction. After compiling a list of programs, each fellowships’ webpage was searched to extract a list of graduated fellows. The name of every person found was inputted into a google search alongside the term “oncology”. From the first 2 pages of results, I attempted to identify their institution for general hematology and oncology fellowship, as well as the institution after their advanced fellowship. I also searched LinkedIn for each fellowship type and reviewed the first 5 pages of people’s results to obtain individuals fitting inclusion-criteria (US-based physicians with previous clinical hematology/oncology fellowship). Fellowships and practicing physicians outside the United States of America who had not completed training in hematology or oncology were excluded, as well as programs specific to pediatrics. As this study concerned only publicly available information, it was not submitted for institutional review board approval.

## Results

14 BMT fellowship programs, 3 lymphoma, 3 leukemia, and 4 Phase 1 fellowships were identified. 6 of the 24 programs listed their alumni information. In total, 66 physicians were found and 56 were analyzed after excluding international physicians without previous hematology/oncology fellowship experience. 25 of the 33 BMT fellow physicians studied were from Stanford. 19 of the 23 leukemia fellow physicians were from MD Anderson.

5 physicians performed hematology/oncology fellowship and remained on faculty at the same place as their specialized fellowship. 9 of the 56 (16%) continued their hematology/oncology fellowships at the same institution of their specialized fellowship. 13/56 (23%) remained at the institution as faculty or attending after their fellowship. 38 (68%) included their specialization in their current position. As shown in the **Figure**, 30% (17/56) of fellows either trained at the same institution or remained at the same institution.

**Figure.**
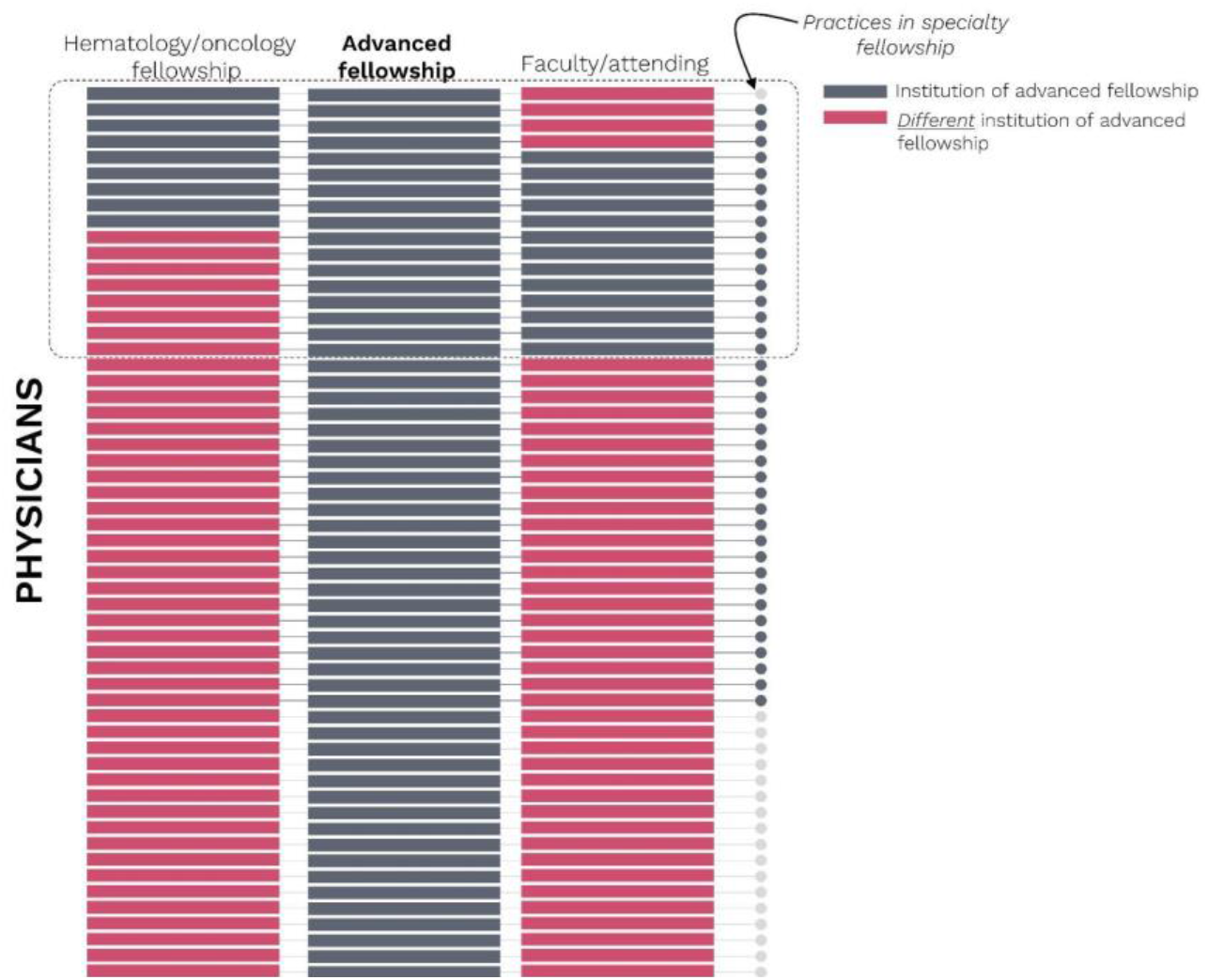
Type of institution after hematology/oncology fellowship for individual physicians studied (n=56)

## Discussion

With a growth in fellowship programs, I sought to examine the empirical characteristics of this new type of training. Through public information, 56 people took part in such programs, of which 30% either trained at that institution or remained there. This raises the question– for at least 1 in 3 physicians– why they had to accept lower salaries and prolonged training when they could have learned these skills during the prior fellowship, or on the job, in the future position.

BMT and other hematologic malignancies are complicated fields which require specialized knowledge, but the best source for such niche learnings are unclear. For example, procedures in bone marrow transplants have been ongoing since the 1980s, but the designated BMT fellowship is a seemingly more recent development.^7^ Historically, BMTs were learned through a combination of hematology fellowship and on-the-job training of attending physicians. BMT fellowships and similar hematologic malignancies programs take a new approach by offering a packaged educational program, circumventing this prior patchwork system but lengthening the medical training process further. This study, which looked into these fellowships, provides no definitive conclusions, but raises the question of whether these fellowships are optimized for the physician’s best-interest.

There are several downsides to prolonging training. Data show that longer work hours decrease the number of women in a field, and long years of training may displace promising people from medicine.^8^ Longer training provides greater opportunity for loan balances to grow and further delays the start of independent clinical practice, which is already now routinely delayed till one’s 30s or 40s. Fellowships may provide didactic instruction but also extract low cost labor. Fellows generally earn far less than similar attending physicians. It is difficult to know to what degree the rise of these programs reflects a genuine educational need versus a corporate-driven convenience.

Justifications for specialized fellowships and the lengthening of medical training are not without possible merit. A recent survey of alumni of BMT fellowship programs who chose to complete the survey, reported a subjective benefit from their training.^9^ Unfortunately, this survey was unable to quantify the opinions and frequency of those who did not respond, and, moreover, what these respondents would say had they not taken an extra year. As such, it provides little usable data.

Advancements in medicine, an increase in number of patients, and a greater reliance on business-centric practices has resulted in physicians requiring more knowledge than previous generations. Informal, on-the-job training can also be more susceptible to uneven workplace dynamics, creating less uniformed training and leaving physicians from underrepresented groups with varying opportunities to obtain skills.^10,11^ However, when adding more years to an already time-intensive process, we should carefully consider the best-use of implementation and the best value added to patient care. For fellows already at an institution, or those joining it for years on end, the use of these fellowships may be debated.

### Limitations

The selection of advanced fellowships was based on individual, non-evidence academic clinician’s experience, and thus susceptible to personal bias. Lack of centralized data for both programs and individuals limited the amount of the information abstracted, resulting in a large amount of missing data. Missing data were also skewed since Stanford BMT and MD Anderson leukemia fellowships had the most extensive public information available, making it the majority of the research sample. At the same time, these are prestigious institutions, where extra fellowship years may be sought. Better public reporting and future research with more complete data is desirable. This research is also specific to the United States of America’s structure of medical training, as well as adult-focused programs.

## Conclusion

We found 1 in 3 participants in additional hematologic malignancy and bone marrow transplant fellows either trained at that institution or remained at that institution, raising the question of whether this dedicated year of training was necessary for their career development, or, if instead, it could have been built in before or after, increasing career earnings. The growth of medical training and unintended consequences should be considered.

## Data Availability

All data produced in the present study are available upon reasonable request to the authors

## Acknowledgements

Jordan Tuia, BA for assistance with figure preparation and data collection.

